# Impact of Providing Future Cardiovascular Risk Based on Genetic Testing on Low-Density Lipoprotein Cholesterol in Patients with Familial Hypercholesterolemia (GenTLe-FH): A Randomized Wait-list Controlled Open-Label Trial

**DOI:** 10.1101/2023.03.26.23287767

**Authors:** Akihiro Nomura, Hirofumi Okada, Atsushi Nohara, Masaaki Kawashiri, Masayuki Takamura, Hayato Tada

## Abstract

**Background and Aims:** Familial hypercholesterolemia (FH) is an autosomal dominant monogenic disease characterized by high low-density lipoprotein cholesterol (LDL-C) levels. Although carrying causative FH variants is associated with coronary heart disease (CHD), it remains unclear whether disclosing its associated cardiovascular risk affects outcomes in patients with FH. Here, we evaluated the efficacy of providing future cardiovascular risk based on genetic testing in addition to a standard FH education program.

**Methods:** We conducted a randomized, wait-list controlled, open-label, single-center trial. In the intervention group, we reported a future cardiovascular risk based on the genetic testing adding to standard FH education at week 0. In the wait-list control group, we only disseminated standard FH education according to the guidelines at week 0; they later received a genetic testing-based cardiovascular risk assessment at week 24. The primary endpoint of this study was the plasma LDL-C level at week 24.

**Results:** Fifty eligible patients with clinically diagnosed FH, without a history of CHD, were allocated to the intervention group (n=24) or the wait-list control group (n=26). At week 24, the intervention group had a significantly greater reduction in LDL-C levels than the wait-list control group (mean changes, -13.1 mg/dL vs. 6.6 mg/dL; difference, -19.7 mg/dL; 95% confidence interval, -34 to -5.6; p=0.009). This interventional effect was consistent with FH causative variant carriers but not with non-carriers.

**Conclusions:** In addition to standard FH care, providing future cardiovascular risk based on genetic testing can further reduce plasma LDL-C levels, particularly among FH causal variant carriers.

**Registration:** Japan Registry of Clinical Trials (jRCTs04218002). URL: https://jrct.niph.go.jp/latest-detail/jRCTs042180027

## Introduction

Familial hypercholesterolemia (FH) is an autosomal dominant Mendelian disease, one of the leading causes of premature coronary heart disease (CHD) along with life-long exposure to high low-density lipoprotein (LDL) cholesterolemia. ^1-6^ The prevalence of patients with heterozygous FH is nearly 0.2-0.5% of the general population (1 in 200-500 individuals). Major FH causative genes are LDL receptor (*LDLR*), proprotein convertase subtilisin/kexin type 9 (*PCSK9*), and apolipoprotein B (*APOB*) and LDLR adaptor protein 1 (*LDLRAP1*, for a particular case of autosomal recessive hypercholesterolemia). FH’s clinical guidelines of FH across the world recommend that patients with FH should be treated using lipid-lowering agents at a younger age to reduce plasma LDL cholesterol (LDL-C) levels, which can prevent future episodes of CHD. Therefore, it is important to diagnose FH as early as possible.^7^

Because FH is a monogenic disorder, genetic testing is important for a definitive diagnosis.^8^ Additionally, our previous study and other studies showed that FH-related pathogenic variants are significantly associated with a higher risk of CHD,^9,10^ which strongly suggests that genetic testing can be used for risk stratification. However, it remains unclear whether FH-related genetic testing would have a potential to affects patients’ outcomes beyond risk stratification. Recently, Kullo et al. reported that disclosure of CHD risk estimates with polygenic risk information, which consists of multiple small-effect genome-wide common genetic variations, led to lower LDL-C levels than disclosure of CHD risk based on classical risk factors alone.^11^ Thus, does providing cardiovascular risk based on FH-related genetic testing also have an impact on changing outcomes in patients with FH?

Here, we examined whether informing future cardiovascular risk based on genetic testing besides conventional FH patient education leads to reduced LDL-C levels in patients with FH.

## Methods

### Overall trial design

This study was a randomized, wait-list controlled, open-label, single-center trial. The detailed trial protocol had been published elsewhere.^8^ In brief, we performed genetic counseling and informed individuals about future cardiovascular risk based on patients’ monogenetic testing results for the intervention group. The primary outcome was the change in plasma LDL-C levels at 24 weeks from the baseline. This trial was conducted in compliance with the Declaration of Helsinki, Ethical Guidelines for Medical and Health Research Involving Human Subjects, and all other applicable laws and guidelines in Japan. The study protocol was approved by the Institutional Review Board of Kanazawa University Hospital (Kanazawa, Japan) and registered in the Japan Registry of Clinical Trials (jRCTs042180027).

### Participants

We recruited patients with clinically diagnosed FH from March 2018 to March 2020 and followed them up until October 2021. We enrolled participants who met all the following inclusion criteria: 1) age ≥15 years; 2) clinically diagnosed with FH according to the Japan Atherosclerosis Society guidelines^1,9^ (if two of the three following criteria were met: [i] LDL-C ≥180 mg/dL, [ii] presence of tendon xanthomas; and [iii] family history of FH or premature coronary artery disease); and 3) never had a genetic test or had not returned genetic results regarding FH. We also excluded participants who met any of the following exclusion criteria: 1) liver dysfunction (aspartate aminotransferase or alanine aminotransferase >3 times the upper normal limits); 2) renal dysfunction (serum creatinine ≥2.0 mg/dL); 3) immunosuppressive state; 4) active cancer; 5) history of CHD including myocardial infarction, percutaneous coronary intervention or coronary artery bypass graft, or coronary artery stenosis (≥75%) previously detected by coronary angiography; or 6) females expecting to be pregnant or currently pregnant. Written informed consent was obtained from all trial participants.

### Screening of FH pathogenic variants

We sequenced the exons of four FH-related genes (*LDLR, PCSK9, APOB*, and autosomal recessive inheritance of *LDLRAP1*) using Illumina MiSeq (San Diego, USA). We defined variants as FH causal when they matched any of the following criteria: a) registered as pathogenic/likely pathogenic in the ClinVar database; b) minor allele frequency <1% in the East Asian population with i) protein-truncating variants (nonsense, canonical splice sites, or frameshift) or ii) missense variants with five *in silico* damaging scores (SIFT, PolyPhen-2 HDIV, PolyPhen-2 HVAR, MutationTaster2, LRT), all predicted as pathogenic;^10^ c) missense variants reported as pathogenic in the Japanese population: *PCSK9* p.Val4Ile and p.Glu32Lys;^11,12^ and d) predicted by eXome-Hidden Markov Model (XHMM) software as copy number variations (large duplication/large deletion).^13^

### Intervention and wait-list control

We randomized patients to either the intervention or the wait-list control groups with FH using an independent web-based randomization system that included a minimization algorithm balanced for age (≥50 years and <50 years), sex (male and female), and causative variant (positive or negative).

In the intervention group, we performed genetic counseling and informed the patients’ future cardiovascular risk based on the results of their FH-related genetic testing in addition to standard FH patient education. The genetic counseling provided by a qualified physician in clinical genetics consisted of the following components: a) genetic diagnosis; b) outline of FH; c) examining the family history of hyperlipidemia/cardiovascular diseases; d) informing that the results of genetic testing could facilitate research in this field; and e) explaining the physical/mental support system in the hospital. In addition, primary cardiologists provided odds ratios of future cardiovascular risk based on the presence or absence of 1) a causal genetic variant and 2) a clinical sign (xanthomas and/or family history of FH) using the original Japanese documents.^8,10,14^ After counseling, we set sufficient time to answer the questions from the patients, confirming their level of understanding.

In the wail-list control group, we only disseminated standard FH patient education using the Japanese booklet for FH patient education according to the FH management guideline.^1^ After the education session, we set the time to answer the queries from the patients. After evaluating the primary endpoint (24^th^ week after randomization), the wait-list control group also received their genetic testing results and future cardiovascular risks via counseling.

In both groups, primary cardiologists provided the standard FH education for their patients. Also, primary cardiologists managed LDL cholesterol levels of their patients at their own discretion according to the Japanese guideline for FH 2017 (targeting LDL cholesterol level of <100 mg/dL or >50% reduction compared with an untreated level for primary prevention).^9^ When adding any of lipid-lowering agents as statins, ezetimibe, or PCSK9 inhibitors, the patients and the physicians must agree with each other for the treatments that were essential to achieve the target value of LDL cholesterol level. Moreover, the number of additional treatments were compared between the intervention and the wait-list control groups. Although in-person FH education was not provided outside of the scheduled outpatient clinic visits, patients in both groups could receive additional counseling and/or outpatient visit when patients wanted or were afraid of genetic testing regardless of the allocated groups during and after the trial.

### Outcomes

The primary outcome was the change in plasma LDL-C levels from baseline (week 0) to week 24. Key secondary outcomes were as follows: 1) changes in plasma LDL-C levels from baseline at week 48 and 2) the Patient Satisfaction Questionnaire Short Form (PSQ-18) scales at weeks 24 and 48. We assessed these outcomes between the intervention and the wait-list control groups or among the four groups categorized by genetic testing results (FH-related causal variant, positive or negative) ([1] intervention + positive; [2] intervention + negative; [3] wait-list control + positive; and [4] wait-list control + negative).

The PSQ-18 scale evaluates patients’ inner satisfaction level for the medical care they received from seven aspects.^15^ We used this scale because providing future cardiovascular risk based on genetic testing could create stress for the patients in terms of longer consultation time, higher medical costs, and more drug prescriptions to achieve the target cholesterol level, particularly for the intervention group. The scale values from 1 to 5 represent the lowest to highest rating for each aspect.

### Statistical analysis and early termination due to the coronavirus disease 19 (COVID-19) pandemic

We hypothesized that the estimated difference from baseline to week 24 in LDL-C levels between the intervention and the wait-list control groups would be 15 mg/dL, with a standard deviation of 25. In the sample size calculation, 44 patients per group were required, with a two-sided α of 5% and power of 80%. We then set the drop-out rate to 10% and determined that approximately 100 patients with FH were required as initially planned.^8^ However, we had to stop the trial enrollment in April 2020 due to the COVID-19 pandemic, with a state of emergency in Japan during the enrollment period. The principal investigator and sub-investigators all agreed to the decision, and we closed the trial in November 2021 after all patients who agreed to enroll in the trial completed the protocol schedule before trial termination.

We compared the outcomes between the intervention and the wait-list control groups. The baseline profiles were described by mean and standard deviation, median and quantiles (continuous variables), or proportion (categorical variables). In addition, we assessed the primary outcome based on the intention-to-treat approach, which was compared between groups using the *t*-test. Moreover, we compared the secondary endpoints between the groups at each defined period using the *t*-test, Mann-Whitney *U*-test, Fisher’s exact test, or linear or logistic regression adjusted for appropriate covariates. A p-value < 0.05 was considered statistically significant for the primary outcome. Statistical analyses were performed using R software version 4.1.2 or above (The R Project for Statistical Computing, Vienna, Austria).

## Results

**Figure 1** shows the flowchart of the trial. We recruited 53 clinically diagnosed FH patients without a history of cardiovascular disease from March 2018 to March 2020 and followed them up until October 2021. Of these, three patients were excluded from the trial because of inappropriate eligibility (n=2) and randomization violation (n=1). The remaining 50 eligible patients were randomly allocated to the intervention group (n=24) or the wait-list control group (n=26). In the intention-to-treat principle, we included all 50 patients for further analyses. The mean age of the patients was 52 ± 16 years, 58% were female, 22% had hypertension, and 8% had diabetes mellitus. Regarding FH-related profiles, 76% had Achilles tendon xanthomas, 60% reported a family history of FH, and the mean plasma LDL-C level was 127 ± 25 mg/dL. The number of FH-related causative genetic variant positive patients was 34 (68%): 31 patients had causal variants at *LDLR* gene, 1 had causal variant (p.Glu32Lys) at *PCSK9* gene, and 2 had copy number variations at *LDLR* gene. We did not detect any causal variants at *APOB* or *LDLRAP1*. The baseline characteristics were well balanced between the intervention and the wait-list control groups (**Table 1**). Since one patient in the wait-list control group refused to follow scheduled visits and laboratory tests after randomization, the study investigators excluded the patient from the study and following outcome analyses.

**Table 1.** Baseline characteristics.

|  | Total | Intervention | Wait-list control |
| --- | --- | --- | --- |
| **N** | 50 | 24 | 26 |
| **Age, years** | 52 ± 16 | 54 ± 15 | 50 ± 17 |
| **Female, n (%)** | 29 (58) | 13 (54) | 16 (62) |
| **Body weight, kg** | 64.0 [55-69] | 64.0 [55-68] | 64.1 [55-71] |
| **BMI, kg/m<sup>2</sup>** | 23.4 [22-27] | 23.7 [21-28] | 23.0 [22-26] |
| **Systolic blood pressure, mmHg** | 116 ± 16 | 116 ± 14 | 117 ± 17 |
| **Diastolic blood pressure, mmHg** | 70 ± 10 | 70 ± 10 | 70 ± 11 |
| **Heart rate, bpm** | 67 ± 11 | 66 ± 13 | 69 ± 9 |
| **Comorbidities, n (%)** |  |  |  |
| Hypertension | 11 (22) | 6 (25) | 5 (19) |
| Diabetes mellitus | 4 (8) | 1 (4) | 3 (12) |
| Current smoking | 5 (10) | 2 (8) | 3 (12) |
| **FH-related profile, n (%)** |  |  |  |
| Achilles tendon xanthomas | 38 (76) | 19 (79) | 19 (73) |
| Family history of FH | 30 (60) | 12 (50) | 18 (69) |
| **Lipid-lowering agents at baseline, n (%)** |  |  |  |
| Statins | 48 (96) | 23 (96) | 25 (96) |
| Ezetimibe | 34 (68) | 17 (71) | 17 (65) |
| PCSK9 inhibitors | 1 (2) | 0 | 1 (4) |
| **Genetic variants, n (%)** |  |  |  |
| Positive | 34 (68) | 17 (71) | 17 (65) |
| At *LDLR* gene | 31 | 15 | 16 |
| At *PCSK9* gene | 1 | 1 | 0 |
| At *APOB* gene | 0 | 0 | 0 |
| At *LDLRAP1* gene (AR inheritance) | 0 | 0 | 0 |
| Copy number variations (at *LDLR* gene) | 2 | 1 | 1 |
| Negative | 16 (32) | 7 (29) | 9 (35) |
| **Lipid profiles** |  |  |  |
| Total cholesterol, mg/dL | 205 ± 31 | 209 ± 33 | 200 ± 29 |
| Triglycerides, mg/dL | 100 ± 65 | 112 ± 74 | 90 ± 55 |
| High-density lipoprotein cholesterol, mg/dL | 59 ± 13 | 57 ± 12 | 60 ± 14 |
| Low-density lipoprotein cholesterol, mg/dL | 127 ± 25 | 133 ± 28 | 121 ± 21 |
| **PSQ-18 scale aspect** |  |  |  |
| General satisfaction | 3.98 ± 0.62 | 3.96 ± 0.67 | 4.00 ± 0.58 |
| Technical quality | 3.99 ± 0.52 | 3.89 ± 0.59 | 4.09 ± 0.45 |
| Interpersonal manner | 4.09 ± 0.57 | 4.08 ± 0.62 | 4.10 ± 0.53 |
| Communication | 4.34 ± 0.41 | 4.38 ± 0.45 | 4.31 ± 0.38 |
| Financial aspects | 3.30 ± 0.68 | 3.21 ± 0.85 | 3.39 ± 0.48 |
| Time spent with a doctor | 3.74 ± 0.61 | 3.85 ± 0.65 | 3.64 ± 0.56 |
| Accessibility and convenience | 3.71 ± 0.49 | 3.73 ± 0.49 | 3.68 ± 0.49 |

Continuous values were expressed as mean ± standard deviation or median (interquartile range). Abbreviations: AR, autosomal recessive; BMI, body mass index; bpm, beats per minutes; FH, familial hypercholesterolemia; LDLR, low-density lipoprotein receptor; PCSK9, proprotein convertase subtilisin/kexin type 9; PSQ-18, Patient Satisfaction Questionnaire Short Form.

**Figure 1.**
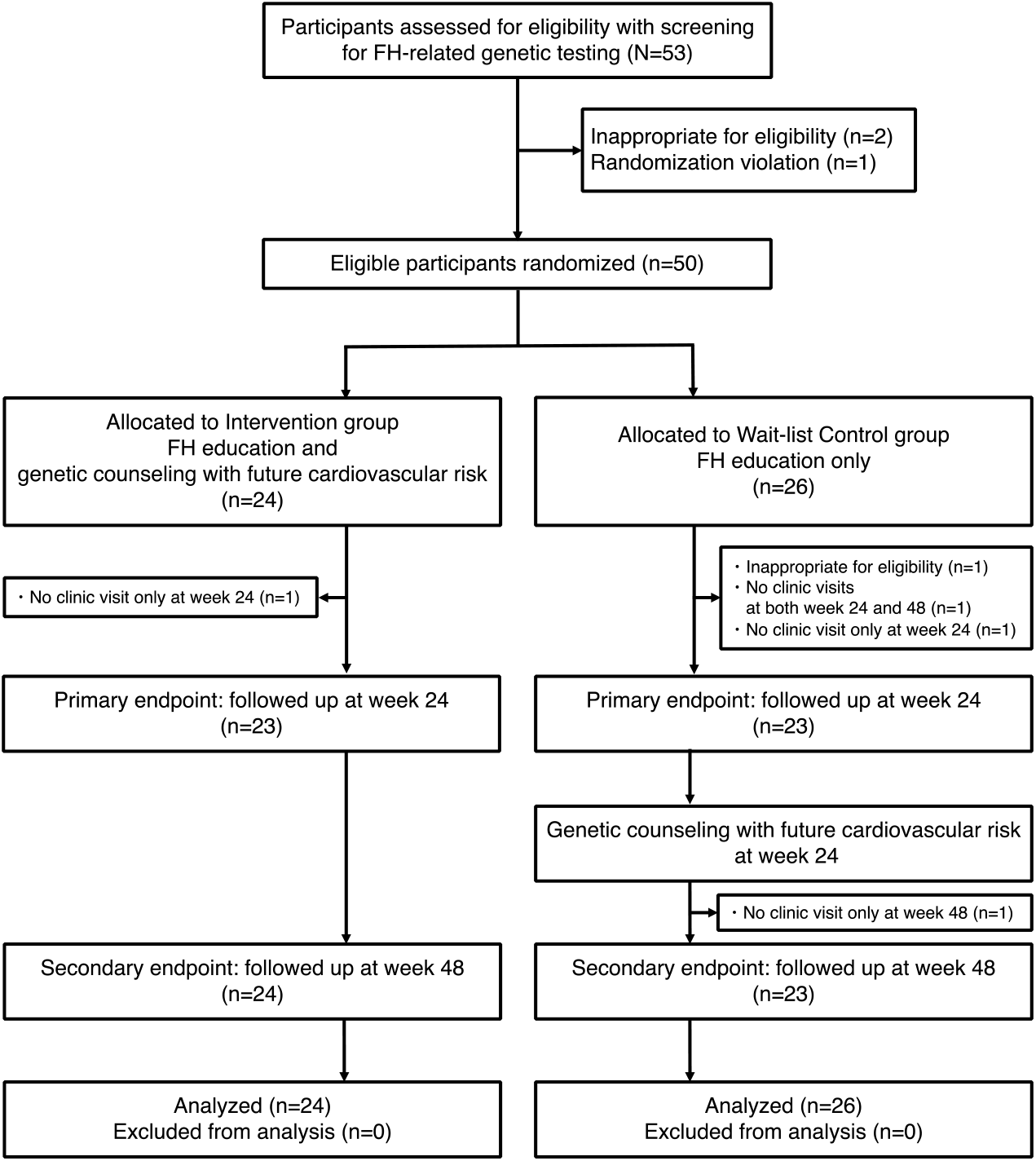
Trial flowchart. We enrolled 53 patients with clinically diagnosed FH and no prior coronary artery disease. Of those, 50 eligible FH patients were randomly allocated to the intervention group (24 patients receiving conventional FH education and future cardiovascular risk assessment based on genetic testing) or the wait-list control group (26 patients receiving only conventional FH education). We followed the two groups for up to 48 weeks and performed an intention-to-treat analysis. The primary endpoint was the change in LDL-C levels from baseline at week 24. Because we handled the wait-list control group as a wait-list group, we also provided future cardiovascular risk assessment based on genetic testing after evaluating the primary endpoint at week 24.

### Primary outcome

At week 24, the intervention group had a significantly greater reduction in plasma LDL-C levels than the wait-list control group (mean changes from baseline, -13.1 mg/dL vs. 6.6 mg/dL; between-group difference, -19.7 mg/dL; 95% confidence interval [CI], -34 to -5.6; p=0.009) (**Table 2, Figure 2**). Even when adjusted by the baseline LDL-C levels, the result was consistent (adjusted difference, -18.0 mg/dL; 95% CI, -32 to -3.8; p=0.02).

**Table 2.** Within-group changes and between-group differences of LDL cholesterol levels.

|  | **week 0 (baseline)**<br>**vs.**<br>**week 24** | **week 24**<br>**vs.**<br>**week 48** | **week 0 (baseline)**<br>**vs.**<br>**week 48** |
| --- | --- | --- | --- |
| **1. All participants** |  |  |  |
| **Within-group changes** |  |  |  |
| Intervention group |  |  |  |
| LDL-C, mean change (SE) | -13.1 (6.2) | 4.3 (2.6) | 0.7 (11.3) |
| Wait-list control group |  |  |  |
| LDL-C, mean change (SE) | 6.6 (6.7) | -8.0 (7.0) | -2.0 (9.0) |
| **Between-group differences\*** |  |  |  |
| LDL-C, mean change [95% CI] | -19.7<br>[-33.8 to -5.6]<br>p=0.009** | 12.3<br>[-2.0 to 26.6]<br>p=0.10 | 2.7<br>[-25.6 to 31.1]<br>p=0.85 |
| **2. Causative variant positive** |  |  |  |
| **Within-group changes** |  |  |  |
| Intervention group |  |  |  |
| LDL-C, mean change (SE) | -19.8 (6.6) | 6.0 (3.0) | -0.1 (15.6) |
| Wait-list control group |  |  |  |
| LDL-C, mean change (SE) | 2.9 (5.6) | -9.6 (11.5) | -7.7 (13.0) |
| **Between-group differences\*** |  |  |  |
| LDL-C, mean change [95% CI] | -22.7<br>[-39.9 to -5.5]<br>p=0.02** | 15.5<br>[-5.7 to 36.7]<br>p=0.16 | 7.6<br>[-33.3 to 48.5]<br>p=0.72 |
| **3. Causative variant negative** |  |  |  |
| **Within-group changes** |  |  |  |
| Intervention group |  |  |  |
| LDL-C, mean change (SE) | 2.3 (6.5) | 0.5 (4.9) | 2.8 (9.1) |
| Wait-list control group |  |  |  |
| LDL-C, mean change (SE) | 12.4 (8.7) | -5.7 (5.1) | 6.8 (11.2) |
| **Between-group differences\*** |  |  |  |
| LDL-C, mean change [95% CI] | -10.1<br>[-32.5 to 12.3]<br>p=0.39 | 6.2<br>[-8.0 to 20.3]<br>p=0.41 | -3.9<br>[-33.4 to 25.6]<br>p=0.80 |

\*:P values were calculated comparing the intervention group with the wait-list control group using the *t*-test. \*\*: p<0.05.

**Figure 2.**
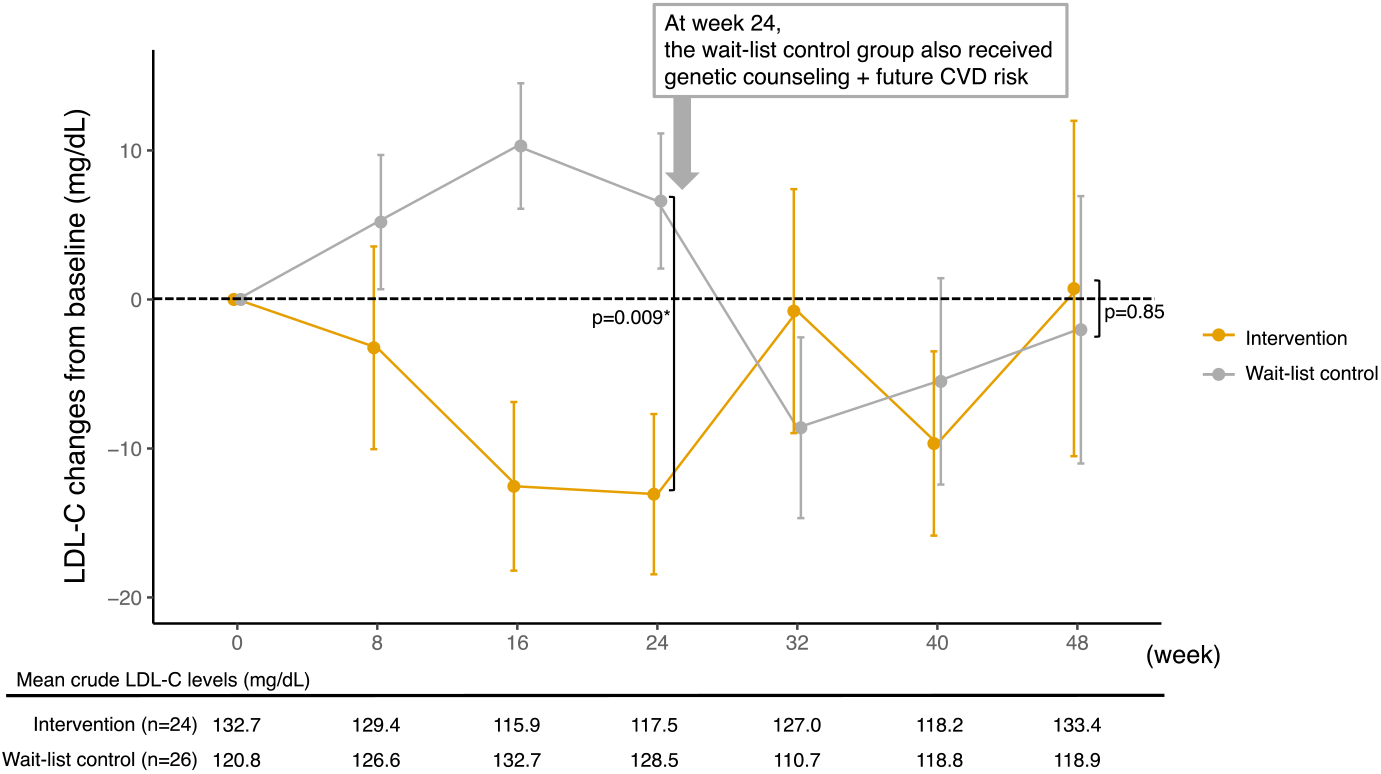
Low-density lipoprotein cholesterol changes from baseline. Regarding the primary endpoint at week 24, the intervention group had a significantly greater reduction in plasma LDL-C level than the wait-list control group. After the wait-list control group also received genetic testing-based future cardiovascular risk assessment at week 24, the difference between the two groups was attenuated at week 48. *:p<0.05. Error bars indicated the standard error.

### Secondary outcomes

After the wait-list control group also received genetic testing-based future cardiovascular risk assessment at week 24, the LDL-C level in the wait-list control group was certainly decreased (the within-group change from week 24 to 48 was -8.0 mg/dL). In contrast, the within-group change in the intervention group from week 24 to 48 was increased by 4.3 mg/dL, that lead to diminish the effect of LDL-C reduction in the intervention group at week 48 from baseline. As a result, the difference between the two groups (between-group difference) from baseline to week 48 was attenuated (0.7 vs. -2.0; between-group difference, 2.8; 95% CI, -26 to 31; p=0.85) (**Table 2**).

**Figure 3** shows the LDL-C changes according to FH-related causative variant carrier status. Among variant carriers, the intervention group also had a greater plasma LDL-C level reduction than the wait-list control group at week 24 (−19.8 mg/dL vs. 2.9 mg/dL; between-group difference, -22.7 mg/dL; 95% CI, -40 to -5.5; p=0.02). However, we could not detect any significant differences between the groups for non-carriers at week 24 (2.3 mg/dL vs. 12.4 mg/dL; between-group difference, -10.1 mg/dL; 95% CI, -33 to 12; p=0.39).

**Figure 3.**
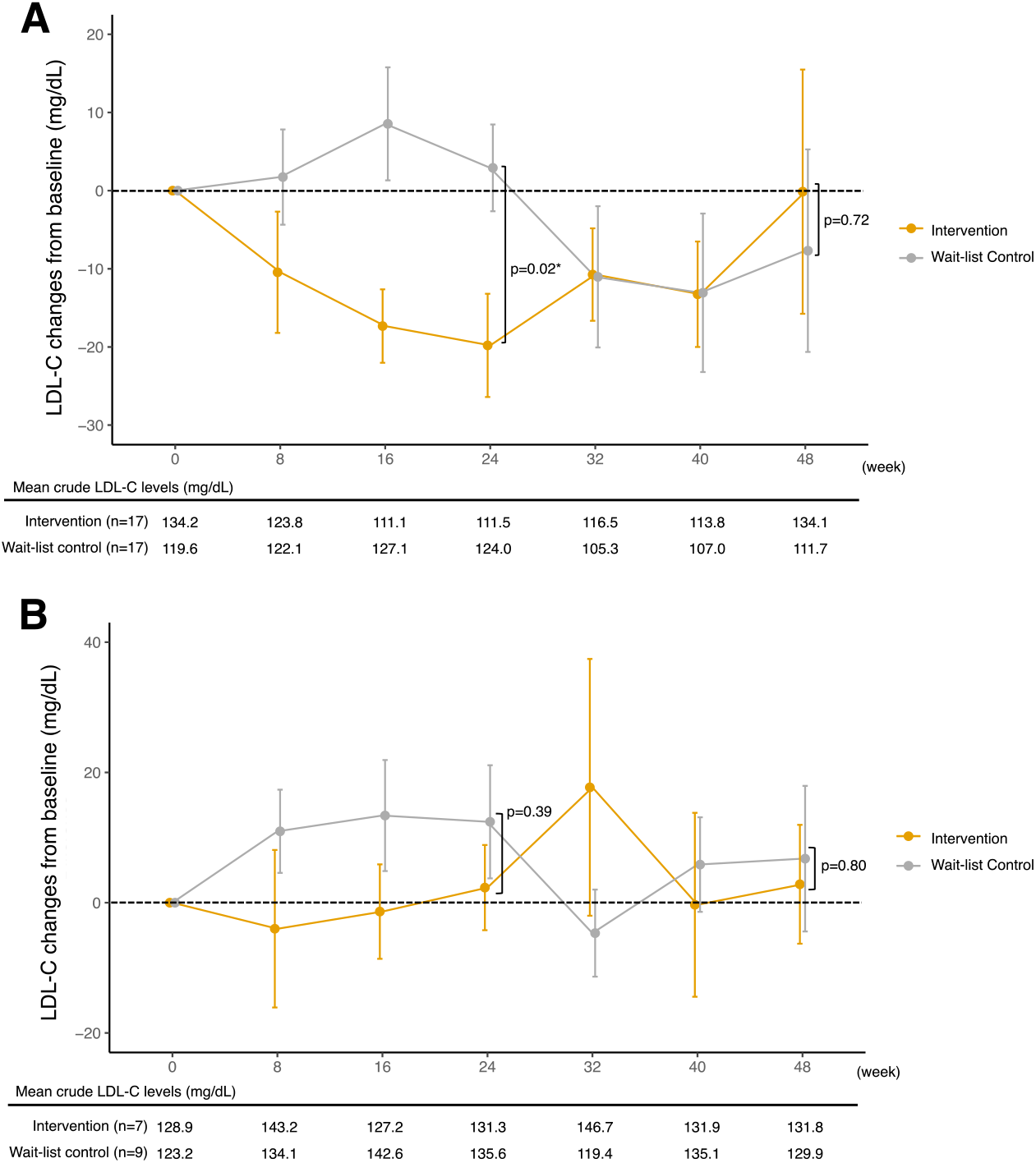
Low-density lipoprotein cholesterol changes from baseline by FH-related causative variant carrier status with additional treatment information. **A**. Variant carriers (n=34). **B**. Non-carriers (n=16). Treatment icons (statin or ezetimibe, statin and ezetimibe, or PCSK9 inhibitor) were placed at the time of each additional medication. Among variant carriers, the intervention group also had a greater plasma LDL-C level reduction than the wait-list control group at week 24. However, we could not detect any apparent differences between the groups for non-carriers. Error bars indicated standard errors

Since the physicians managed LDL cholesterol levels of their patients at their own discretion according to the guideline, we investigated additional treatment regimens on top of the baseline lipid-lowering agents in both groups (**Table 3**). The numbers of participants receiving add-on treatment during the study were 10 (42%) in the intervention group and 6 (23%) in the wait-list control group (chi-square test, p=0.26). Of note, 90% (9/10) in the intervention group and 67% (4/6) in the wait-list control group of add-on lipid-lowering agents were prescribed within 24 weeks from receiving genetic counseling and future cardiovascular risk based on the genetic test. Moreover, the add-on treatments were conducted more frequent in causative variant carrier group (14/16, 88%) than in non-carrier group (2/16, 12%).

**Table 3.**
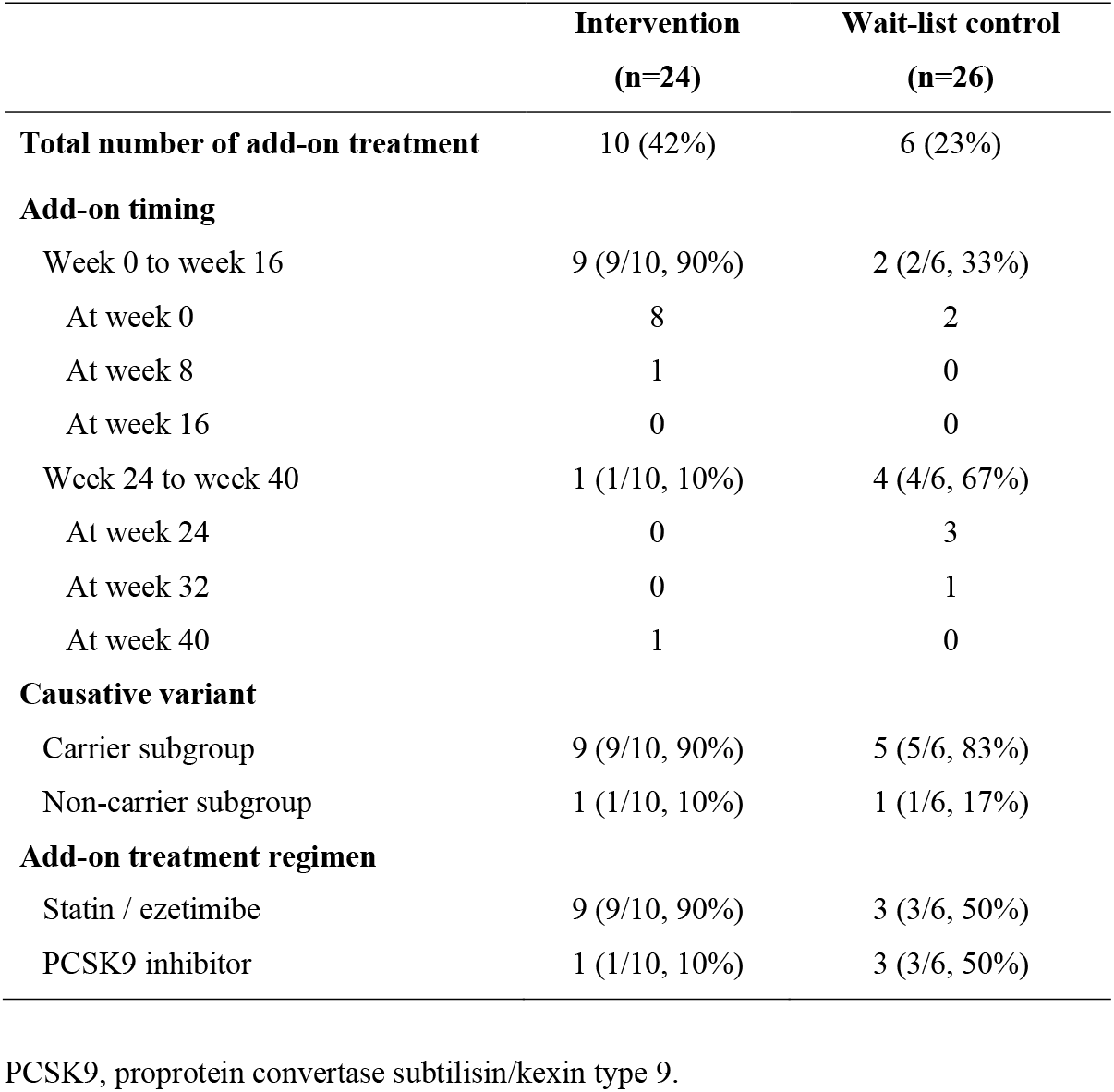
Add-on treatment regimens by primary physicians during study period.

### Patient satisfaction evaluation

We further assessed the FH patients’ satisfaction for medical care they received during the study period by the PSQ-18 scale analysis (**Figure 4**). We evaluated the score changes by seven aspects (general satisfaction, technical quality, interpersonal manner, communication, financial aspects, time spent with doctor, and accessibility and convenience), and there were no between-group differences in each aspect except in the “general satisfaction” aspect for medical care. Notably, FH-related causal variant non-carriers in the intervention group were less satisfied with general medical care than the wait-list control group at 24 weeks.

**Figure 4.**
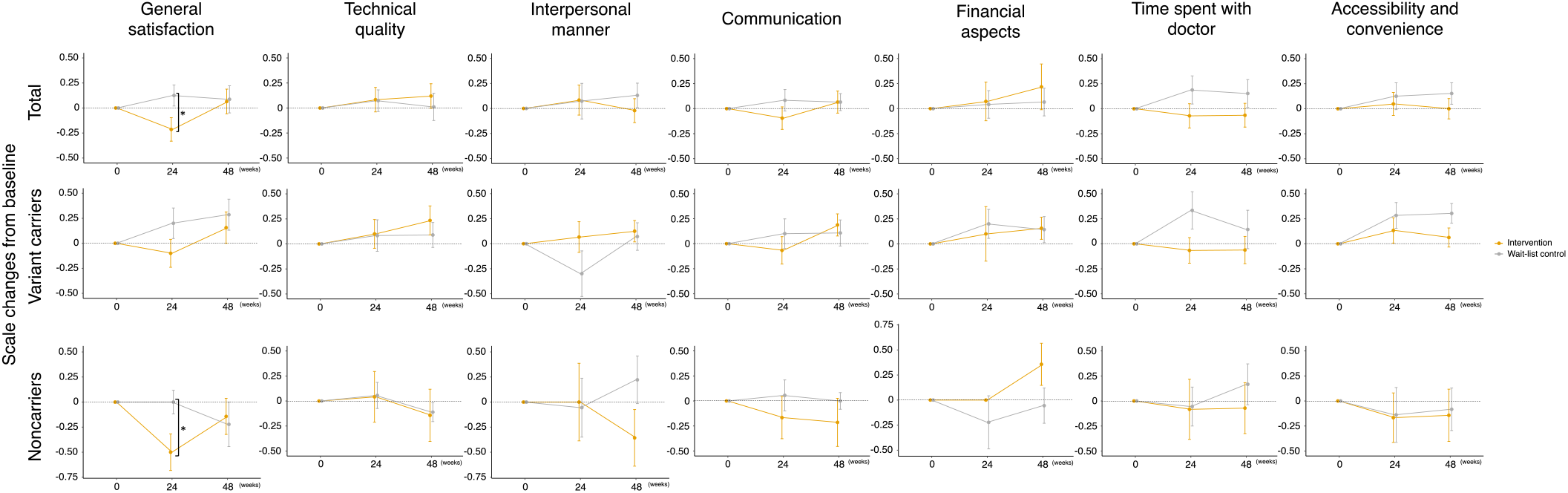
Changes in patient satisfaction questionnaire scale scores by groups. There were no between-group differences except in the “general satisfaction” aspect of medical care. According to the general satisfaction scale results, variant non-carriers in the intervention group were less satisfied with general medical care than the wait-list control group at 24 weeks. *:p<0.05. Error bars indicated the standard error.

### Subgroup analysis

**Table 4** shows the subgroup analysis results for the treatment differences in LDL-C levels at week 24. There were no significant interactions in each subgroup except that age (≥50 vs. <50 years) subgroup was the only significant quantitative interaction between the intervention and the wait-list control groups.

**Table 4.** Treatment differences in low-density lipoprotein cholesterol changes at week 24 between the intervention and the wait-list control groups by patient subgroups.

|  | **Intervention** | **Wait-list control** | **Difference (95% CI)** | **P interaction** |
| --- | --- | --- | --- | --- |
| Age ≥50 (n=26) | -5.2 | 1.4 | -6.6 [-20 to 7.0] | 0.038 |
| Age <50 (n=24) | -23.3 | 13.4 | -36.7 [-62 to -11] |  |
| Male (n=21) | -12.2 | -0.4 | -11.8 [-33 to 9.7] | 0.41 |
| Female (n=29) | -13.8 | 10.4 | -24.2 [-43 to -5.0] |  |
| BMI ≥25 (n=15) | -19.1 | 2.3 | -21.4 [-48 to 5.1] | 0.88 |
| BMI <25 (n=35) | -10.4 | 8.6 | -19.0 [-36 to -2.0] |  |
| Hypertension+ (n=11) | -6.6 | -0.6 | -6.0 [-28 to 16] | 0.30 |
| Hypertension- (n=39) | -15.4 | 8.6 | -24.0 [-41 to -6.7] |  |
| Xanthomas+ (n=39) | -9.5 | 7.7 | -17.2 [-33 to -1.1] | 0.51 |
| Xanthomas- (n=11) | -25.9 | 3.0 | -28.9 [-58 to 0.3] |  |
| Family history of FH+ (n=30) | -12.7 | 8.9 | -21.5 [-39 to -3.6] | 0.71 |
| Family history of FH- (n=20) | -13.5 | 2.4 | -15.9 [-40 to 8.2] |  |
| Causative variant positive (n=34) | -19.8 | 2.9 | -22.7 [-40 to -9.6] | 0.40 |
| Causative variant negative (n=16) | 2.3 | 12.4 | -10.1 [-33 to 12] |  |
| Baseline LDL-C ≥130 (n=21) | -25.8 | 3.4 | -29.2 [-54 to -4.7] | 0.24 |
| Baseline LDL-C <130 (n=29) | -3.3 | 9.1 | -12.4 [-27 to 2.6] |  |
| Adding lipid-lowering agents at week 0 to 16+ (n=11) | -20.4 | 5.4 | -25.8 [-80 to 28] | 0.72 |
| Adding lipid-lowering agents at week 0 to 16- (n=39) | -9.2 | 6.7 | -15.9 [-32 to 0.1] |  |

Abbreviations: BMI, body mass index; CI, confidence interval; FH, familial hypercholesterolemia; LDL-C, low-density lipoprotein cholesterol.

## Discussion

This trial was the first randomized, wait-list controlled study to assess whether disclosing the risk for future cardiovascular diseases based on genetic testing results in addition to standard FH education would lead to reduced LDL-C levels in patients with FH. For the primary endpoint, the intervention group showed a significantly greater reduction in plasma LDL-C levels than the wait-list control group at week 24. The difference between the groups was attenuated at week 48 after the wait-list control group also received future cardiovascular risk assessment based on genetic testing at week 24. On the other hand, as for patient satisfaction evaluation by the PSQ-18, no between-group differences were found except for the general satisfaction aspect of medical care. Notably, non-carriers of FH-related causal variants in the intervention group were less satisfied with general medical care than those in the wait-list control group at 24 weeks.

The conclusions of this study are as follows. First, providing cardiovascular risk based on genetic testing further reduced plasma LDL-C levels, in addition to standard FH care. One of the reasons for the significant reduction effect could be that patients with FH may become aware of the necessity of medication, leading to better adherence to medications and achieving lifestyle modifications by themselves. However, there was a concern that the LDL-C reduction might be derived from more intensive treatments based on the information on genetic testing-based cardiovascular risk. Previous studies reported that providing genetic testing results for primary physicians did not affect their practice because they thought plasma lipid levels were simply sufficient to select treatment options for patients with FH and felt little need for genetic testing.^16,17^ Nevertheless, another studies demonstrated that FH genetic counseling for patients with FH might convince them of the use of more intensified lipid-lowering drugs and improve medication adherence, leading to lower LDL-C levels, irrespective of their treatment status.^18-20^ Moreover, a genetic diagnosis itself could promote the selection of more aggressive lipid-lowering drugs for primary physicians.^21^ For example, the MI-GENES clinical trial, which demonstrated the effect of disclosure of CHD genetic risk on LDL-C levels for patients with intermediate CHD risk, indicated that the lipid-lowering effects were derived from the initiation of statin medication.^22^ In this study, We did not observe any interactions between baseline plasma LDL-C levels and the addition of lipid-lowering agents between weeks 0 and 24 (**Figure 4**). However, although non-significant, an add-on lipid-lowering treatment was more frequent within 24 weeks from receiving genetic counseling and future cardiovascular risk based on the genetic test (**Table 3**), which might have contributed to the favorable result for LDL-C reduction in the intervention group. In addition, genetic confirmation might help patients with FH reinforce a healthier diet and exercise behaviors.^23^ These multilateral effects could affect the LDL-C levels of FH patients without a history of cardiovascular complications.

Second, we observed inconsistent effects of the intervention according to the variant carrier status on LDL-C levels. For FH-related causal variant carriers, the change in LDL-C levels in the intervention group was greater than that in the wait-list control group at week 24, although this favorable change did not occur in non-carriers. As stated above, providing a “positive” genetic testing result for patients may improve adherence to lipid-lowering medication and permit access to specific treatments, such as the PCSK9 inhibitor.^24^ Moreover, Claassen et al. showed that patients with FH who received their genetic testing results had a higher perceived efficacy of lipid-lowering medication than those without.^25^ In contrast, when the FH causal variant was not identified, the negative genetic testing result might provide mental relief even for at-risk patients,^26^ which could have a literal “negative” effect on non-carrier patients. Additionally, it could cause reduced compliance or motivation in patients.^24^

Third, although the significant LDL-C reduction was achieved after 24 weeks from the disclosure of future cardiovascular risk based on genetic testing, the effect was attenuated after 48 weeks of the intervention. The changes in LDL-C levels from genetic counseling with providing cardiovascular risk assessment to 24 weeks were consistent in both groups (−13.1 mg/dL in the intervention group and -8.0 mg/dL in the wait-list control group), whereas the change in LDL-C to 48 weeks was just +0.7 mg/dL despite performing some additional lipid-lowering treatments on the baseline regimen. Although a long-term health behavior change was certainly challenging,^27,28^ our results implicated that the effect of our “one-time” intervention in an outpatient clinic at week 0 might be effective through 6 months but could be attenuated from then on.

Fourth, genetic counseling for FH did not make patients nervous overall, although non-carriers were less satisfied with general medical care than carriers. In general, patient empowerment is a potential non-clinical benefit of genetic testing. Providing a diagnosis could allow the patient to improve their understanding of the disease’s clinical course, treatment options, and psychological control.^26,29^ Genetic testing sometimes ameliorates uncertainty, stigma, and personal guilt.^30,31^ However, it could be valid only in FH patients with positive genetic testing, not in those with negative genetic testing. Patients sometimes undergo genetic testing because they want to confirm whether they are negative. Clinically diagnosed patients with FH are at a higher risk for cardiovascular disease than patients without FH, regardless of the causative variant carrier status. However, when we provide variant-negative patients with less CHD risk than variant-positive FH patients, they might misunderstand the information, become unexpectedly reassured, and decrease their adherence and lifestyle modification. Care might be taken with these “non-carriers” to provide the current FH education strategy with future cardiovascular risk assessment based on the variant-negative genetic testing results. This is because the counseling provided in this study might potentially provide misinterpretation and less satisfaction for medical care to “non-carriers”. Therefore, alternative counseling strategies may be needed for patients with variant-negative FH.

The strength of this study was that it was a randomized trial to demonstrate the efficacy of disclosing future cardiovascular disease risk assessment based on genetic testing results in addition to standard FH education for FH patients without a history of cardiovascular disease. The most important limitation of this study was that while we originally planned to enroll approximately 100 FH patients in this study,^8^ we halted enrollment due to the COVID-19 pandemic and closed recruitment before completing the pre-specified protocol schedule. Although the effect difference on LDL-C was larger than expected and we demonstrated the efficacy of the intervention on LDL-C levels, this unique situation needs to be considered with caution for clinical application of our findings. In addition, in terms of LDL-C levels, the mean LDL-C levels at 48 weeks were still >100 mg/dL in both groups, which was above the recommended lipid control threshold (<100 mg/dL) for FH patients as primary prevention. Although clarifying cardiovascular risk for both FH patients and primary physicians could improve the treatment quality,^32^ achieving the target LDL-C level (<100 mg/dL) has still been a challenging goal, as the CASCADE-FH registry reported.^33^ Although this study’s median LDL-C level was lower than that from the registry, we need to focus on the target cholesterol level to reduce the future cardiovascular risk for patients with FH as much as possible. Furthermore, we could not assessed patients’ medication adherence except patient-reported drug non-compliance that might affect the results.

In conclusion, in addition to standard FH care, providing cardiovascular risk assessment based on genetic testing further reduced plasma LDL-C levels further, particularly among patients with FH causal variants carriers. Meanwhile, we might be careful with the non-carriers to provide this risk disclosure strategy based on the variant-negative genetic testing results.

## Data Availability

The data in this trial are available from the corresponding author upon reasonable request.

## Non-standard Abbreviations and Acronyms

APOB: apolipoprotein B
BMI: body mass index
Bpm: beats per minutes
CHD: coronary heart disease
CI: confidence interval
COVID-19: coronavirus disease 19
CVD: cardiovascular disease
FH: familial hypercholesterolemia
LDL: low-density lipoprotein
LDL-C: low-density lipoprotein cholesterol
LDLR: low-density lipoprotein receptor
LDLRAP1: low-density lipoprotein receptor adaptor protein 1
PCSK9: proprotein convertase subtilisin/kexin type 9
PSQ-18: Patient Satisfaction Questionnaire Short Form

## Acknowledgements

We are very thankful to all the participants and staff regarding this trial. We also express our gratitude to Mitsuyo Kusajima, Emi Tamukai, Ryogo Shimizu, Asako Kanadu, and Rika Miyashita as clinical research coordinators; Kenichi Yoshimura, Hideki Ishikawa for randomization and statistical assistance; Yasuhiko Imai, and Jia Yu for clinical data management; and Toshinori Murayama, chairman of the Innovative Clinical Research Center, Kanazawa University for supporting our clinical trial.

## Data availability statement

The data in this trial are available from the corresponding author upon reasonable request.

## References

1. Harada-Shiba M, Arai H, Oikawa S, et al. Guidelines for the management of familial hypercholesterolemia. J Atheroscler Thromb 2012;19:1043-1060. doi:

2. Gidding SS, Champagne MA, de Ferranti SD, et al. The Agenda for Familial Hypercholesterolemia: A Scientific Statement From the American Heart Association. Circulation 2015;132:2167–2192. doi: 10.1161/CIR.0000000000000297

3. Watts GF, Gidding S, Wierzbicki AS, et al. Integrated guidance on the care of familial hypercholesterolaemia from the International FH Foundation. Int J Cardiol 2014;171:309–325. doi: 10.1016/j.ijcard.2013.11.025

4. Nordestgaard BG, Chapman MJ, Humphries SE, et al. Familial hypercholesterolaemia is underdiagnosed and undertreated in the general population: guidance for clinicians to prevent coronary heart disease: consensus statement of the European Atherosclerosis Society. Eur Heart J 2013;34:3478–3490a. doi: 10.1093/eurheartj/eht273

5. Mabuchi H, Nohara A, Noguchi T, et al. Molecular genetic epidemiology of homozygous familial hypercholesterolemia in the Hokuriku district of Japan. Atherosclerosis 2011;214:404–407. doi: 10.1016/j.atherosclerosis.2010.11.005

6. Versmissen J, Oosterveer DM, Yazdanpanah M, et al. Efficacy of statins in familial hypercholesterolaemia: a long term cohort study. BMJ 2008;337:a2423. doi: 10.1136/bmj.a2423

7. Teramoto T, Kobayashi M, Tasaki H, et al. Efficacy and Safety of Alirocumab in Japanese Patients With Heterozygous Familial Hypercholesterolemia or at High Cardiovascular Risk With Hypercholesterolemia Not Adequately Controlled With Statins-ODYSSEY JAPAN Randomized Controlled Trial. Circ J 2016;80:1980–1987. doi: 10.1253/circj.CJ-16-0387

8. Nomura A, Tada H, Okada H, et al. Impact of genetic testing on low-density lipoprotein cholesterol in patients with familial hypercholesterolemia (GenTLe-FH): a randomised waiting list controlled open-label study protocol. BMJ Open 2018;8:e023636. doi: 10.1136/bmjopen-2018-023636

9. Harada-Shiba M, Arai H, Ishigaki Y, et al. Guidelines for Diagnosis and Treatment of Familial Hypercholesterolemia 2017. J Atheroscler Thromb 2018;25:751–770. doi: 10.5551/jat.CR003

10. Khera AV, Won HH, Peloso GM, et al. Diagnostic Yield and Clinical Utility of Sequencing Familial Hypercholesterolemia Genes in Patients With Severe Hypercholesterolemia. J Am Coll Cardiol 2016;67:2578–2589. doi: 10.1016/j.jacc.2016.03.520

11. Mabuchi H, Nohara A, Noguchi T, et al. Genotypic and phenotypic features in homozygous familial hypercholesterolemia caused by proprotein convertase subtilisin/kexin type 9 (PCSK9) gain-of-function mutation. Atherosclerosis 2014;236:54–61. doi: 10.1016/j.atherosclerosis.2014.06.005

12. Ohta N, Hori M, Takahashi A, et al. Proprotein convertase subtilisin/kexin 9 V4I variant with LDLR mutations modifies the phenotype of familial hypercholesterolemia. J Clin Lipidol 2016;10:547–555 e545. doi: 10.1016/j.jacl.2015.12.024

13. Fromer M, Moran JL, Chambert K, et al. Discovery and statistical genotyping of copy-number variation from whole-exome sequencing depth. Am J Hum Genet 2012;91:597–607. doi: 10.1016/j.ajhg.2012.08.005

14. Tada H, Kawashiri MA, Nohara A, et al. Impact of clinical signs and genetic diagnosis of familial hypercholesterolaemia on the prevalence of coronary artery disease in patients with severe hypercholesterolaemia. Eur Heart J 2017. doi: 10.1093/eurheartj/ehx004

15. Marshall G, Hays R. The Patient Satisfaction Questionnaire Short Form (PSQ-18). Santa Monica, CA: RAND Corporation; 1994. p 7865.

16. Will CM, Armstrong D, Marteau TM. Genetic unexceptionalism: clinician accounts of genetic testing for familial hypercholesterolaemia. Soc Sci Med 2010;71:910–917. doi: 10.1016/j.socscimed.2010.05.018

17. Lerner B, Marshall N, Oishi S, et al. The value of genetic testing: beyond clinical utility. Genet Med 2017;19:763–771. doi: 10.1038/gim.2016.186

18. Umans-Eckenhausen MA, Defesche JC, Sijbrands EJ, Scheerder RL, Kastelein JJ. Review of first 5 years of screening for familial hypercholesterolaemia in the Netherlands. Lancet 2001;357:165–168. doi: 10.1016/S0140-6736(00)03587-X

19. Leren TP. Cascade genetic screening for familial hypercholesterolemia. Clin Genet 2004;66:483–487. doi: 10.1111/j.1399-0004.2004.00320.x

20. Sturm AC, Knowles JW, Gidding SS, et al. Clinical Genetic Testing for Familial Hypercholesterolemia: JACC Scientific Expert Panel. J Am Coll Cardiol 2018;72:662–680. doi: 10.1016/j.jacc.2018.05.044

21. Paynter NP, Ridker PM, Chasman DI. Are Genetic Tests for Atherosclerosis Ready for Routine Clinical Use? Circ Res 2016;118:607–619. doi: 10.1161/CIRCRESAHA.115.306360

22. Kullo IJ, Jouni H, Austin EE, et al. Incorporating a Genetic Risk Score Into Coronary Heart Disease Risk Estimates: Effect on Low-Density Lipoprotein Cholesterol Levels (the MI-GENES Clinical Trial). Circulation 2016;133:1181–1188. doi: 10.1161/CIRCULATIONAHA.115.020109

23. Hagger MS, Hardcastle SJ, Hingley C, et al. Predicting Self-Management Behaviors in Familial Hypercholesterolemia Using an Integrated Theoretical Model: the Impact of Beliefs About Illnesses and Beliefs About Behaviors. Int J Behav Med 2016;23:282–294. doi: 10.1007/s12529-015-9531-x

24. Brown EE, Sturm AC, Cuchel M, et al. Genetic testing in dyslipidemia: A scientific statement from the National Lipid Association. J Clin Lipidol 2020;14:398–413. doi: 10.1016/j.jacl.2020.04.011

25. Claassen L, Henneman L, van der Weijden T, Marteau TM, Timmermans DR. Being at risk for cardiovascular disease: perceptions and preventive behavior in people with and without a known genetic predisposition. Psychol Health Med 2012;17:511–521. doi: 10.1080/13548506.2011.644246

26. Severin F, Borry P, Cornel MC, et al. Points to consider for prioritizing clinical genetic testing services: a European consensus process oriented at accountability for reasonableness. Eur J Hum Genet 2015;23:729–735. doi: 10.1038/ejhg.2014.190

27. Middleton KR, Anton SD, Perri MG. Long-Term Adherence to Health Behavior Change. Am J Lifestyle Med 2013;7:395–404. doi: 10.1177/1559827613488867

28. Bouton ME. Why behavior change is difficult to sustain. Prev Med 2014;68:29–36. doi: 10.1016/j.ypmed.2014.06.010

29. Berberich AJ, Hegele RA. The role of genetic testing in dyslipidaemia. Pathology 2019;51:184–192. doi: 10.1016/j.pathol.2018.10.014

30. Senior V, Smith JA, Michie S, Marteau TM. Making sense of risk: an interpretative phenomenological analysis of vulnerability to heart disease. J Health Psychol 2002;7:157–168. doi: 10.1177/1359105302007002455

31. Weiner K, Durrington PN. Patients’ understandings and experiences of familial hypercholesterolemia. Community Genet 2008;11:273–282. doi: 10.1159/000121398

32. Jones LK, Sturm AC, Seaton TL, et al. Barriers, facilitators, and solutions to familial hypercholesterolemia treatment. PLoS One 2020;15:e0244193. doi: 10.1371/journal.pone.0244193

33. deGoma EM, Ahmad ZS, O’Brien EC, et al. Treatment Gaps in Adults With Heterozygous Familial Hypercholesterolemia in the United States: Data From the CASCADE-FH Registry. Circ Cardiovasc Genet 2016;9:240–249. doi: 10.1161/CIRCGENETICS.116.001381

